# How a year of pandemic and related public health measures impacted youth and young adults and the foundations they build upon: Qualitative interviews in Ontario Canada

**DOI:** 10.1101/2024.07.06.24310036

**Authors:** Laurel C. Austin, Makayla Nunes Gomes, Sebastian Chavez, Celina Degano

## Abstract

**Introduction:** During youth and young adult (YYA) years education, employment, relationships with family and friends, and important rituals representing transition to new phases of life are foundations on which personal identity and future well-being are established. These were shaken by the COVID-19 pandemic. We explore how Ontario YYAs and the foundations they build upon (education, employment, relationships, transitional events) were impacted by over a year of pandemic and public health responses to prevent spread.

**Methods:** In-depth semi-structured interviews with 19 Ontario YYAs age 16-21 were conducted during April - June 2021. Reflexive thematic analysis aided by MAXQDA software was used to iteratively engage with data to search for patterns and shared meaning.

**Results:** Thirteen themes were identified, organized into four meta-themes: impacts on self, impacts on foundations (educational, employment, transitional events/rituals), impacts on relationships, and coping responses. Many, especially those living with loved ones believed to risk a fatal outcome from COVID-19, felt the weight of needing to avoid the virus to protect loved ones. YYAs who were in their last year of secondary school in spring 2020 or 2021 missed important transitional endings, e.g., graduation. Those graduating in 2020 and going on to post-secondary school also missed transitional beginnings, e.g., experiencing in-person on-campus higher education classrooms, living in residence, and meeting new friends. Perceived negative impacts on education quality and professional development were common. Virtual learning models and changes to in-person schooling, hastily introduced and evolving over the next year, did not measure up to traditional learning models.

**Conclusions:** All of these impacts took a toll. Respondents routinely volunteered concerns about stress, loneliness, and their mental health. There is need for further research to assess long-term impacts of these experiences, especially among YYAs who had family members at severe risk, and those finishing secondary school in spring of 2020.

## Introduction

### Background

In response to the World Health Organization (WHO)’s declaration of SARS-CoV-2 (aka COVID-19) as a global pandemic on March 11, 2020, governments around the world implemented public health measures to limit viral spread. These included social distancing, school closures, travel restrictions, quarantine, mask mandates, “lockdowns”, and more. Policies and definitions of key terms (e.g. ‘stay-at-home’ vs ‘lockdown’) varied across jurisdictions and over time as the pandemic evolved, with over 13,000 policy announcements across 195 countries in the first few months (1).

During YYA years, important foundations for personal identity and future well-being are established. Friendships, social interactions, and growing independence are key to development of personal identity and intimacy (2–4). Friends and parents help young people work through important decisions (5); friends ease transition to post-secondary education (6). Important life events, e.g., graduation and first year of post-secondary school, mark transitions to adulthood (2,4). Quality of education impacts future education, employment, and well-being (7,8). Apprenticeships, internships, and co-ops offer skill development and expanded networks (9–11).

A natural question then is how were YYAs and these important foundations impacted by the pandemic and related public health responses that were in place for an extended period? COVID-19 was the first global pandemic that lasted for over a year since the Spanish Flu which started in 1918. Thus, research on the impacts of an extended pandemic and public health responses was necessarily in a nascent state during the COVID-19 pandemic. Interview based qualitative research, employed in this study, is especially well suited to rich and deep exploration of new phenomenon, for example, how the pandemic and public health measures impacted YYAs and the foundations they build upon.

From the early months of COVID-19, qualitative studies in South Africa (12), Portugal (13), the US, (14), and the UK (15,16) reported on YYA experiences across a range of topics. A study in India (17) focused on concerns related to mandatory university entrance exams. These studies showed that context matters. For example, YYAs in South Africa were worried about basic necessities (12); students in India were unable to take mandatory exams, meaning university entry was delayed a year (17); secondary students in the US were most concerned about academics and work habits, physical health, and mental health (14).

There is a small but growing body of qualitative research on how the extended pandemic impacted YYAs, including in Canada. Often studies focus on vulnerable or under-represented populations, or on limited topics. For example, Riazi et al., examined impacts of extended pandemic on secondary school life across Canada, finding YYAs felt left out of communications about changes that impacted them, didn’t like structural changes implemented during pandemic, missed important events like graduation, and were concerned about mental health and access to supports (18). Interviews with nine Inuit youth leaders in Nunavut, Canada, an Arctic community, focused on how the pandemic impacted their engagement with I-SPARX, an Inuit specific e-intervention designed to build resilience in mental health (19). Nobel et al (20) interviewed homeless youth about the unique structural and psychosocial challenges they faced during an extended pandemic. Our study was designed to broadly explore how the extended pandemic and public health measures to reduce viral spread impacted YYAs in Ontario Canada, examining impacts to the foundations they build their futures upon: relationships, education, employment, and important transitional events, as well as how they coped with all the changes.

### Ontario context

Public health measures were established at provincial and local levels based on COIVD-19 case numbers, hospital capacity, and more, meaning guidelines varied across Ontario’s 34 public health units at times. Table 1 provides a timeline of key measures through June 2021. Province-wide “stay at home orders” were in effect March - June 2020, Dec 2020 – January 2021, and April 12, 2021 – June 2021; this last period coincided with the time of our interviews. The most populated areas had additional times with stay-at-home orders.^1^ All secondary and post-secondary switched to virtual education in March 2020. Secondary schools varied approaches to learning during the 2020-2021 school year (virtual, in-person, hybrid), implemented modified schedules (quad-mesters, staggered lunch), and varied policies (e.g. regarding extracurricular activities) (21). Schools shifted between approaches as case counts and public health measures varied. Ontario led in Canada as the province with the most weeks of in-person school closures due to COVID-19 (22). Post-secondary remained almost entirely online with few exceptions, e.g., for lab work, through August 2021, and longer for some institutions.

**Table 1:**
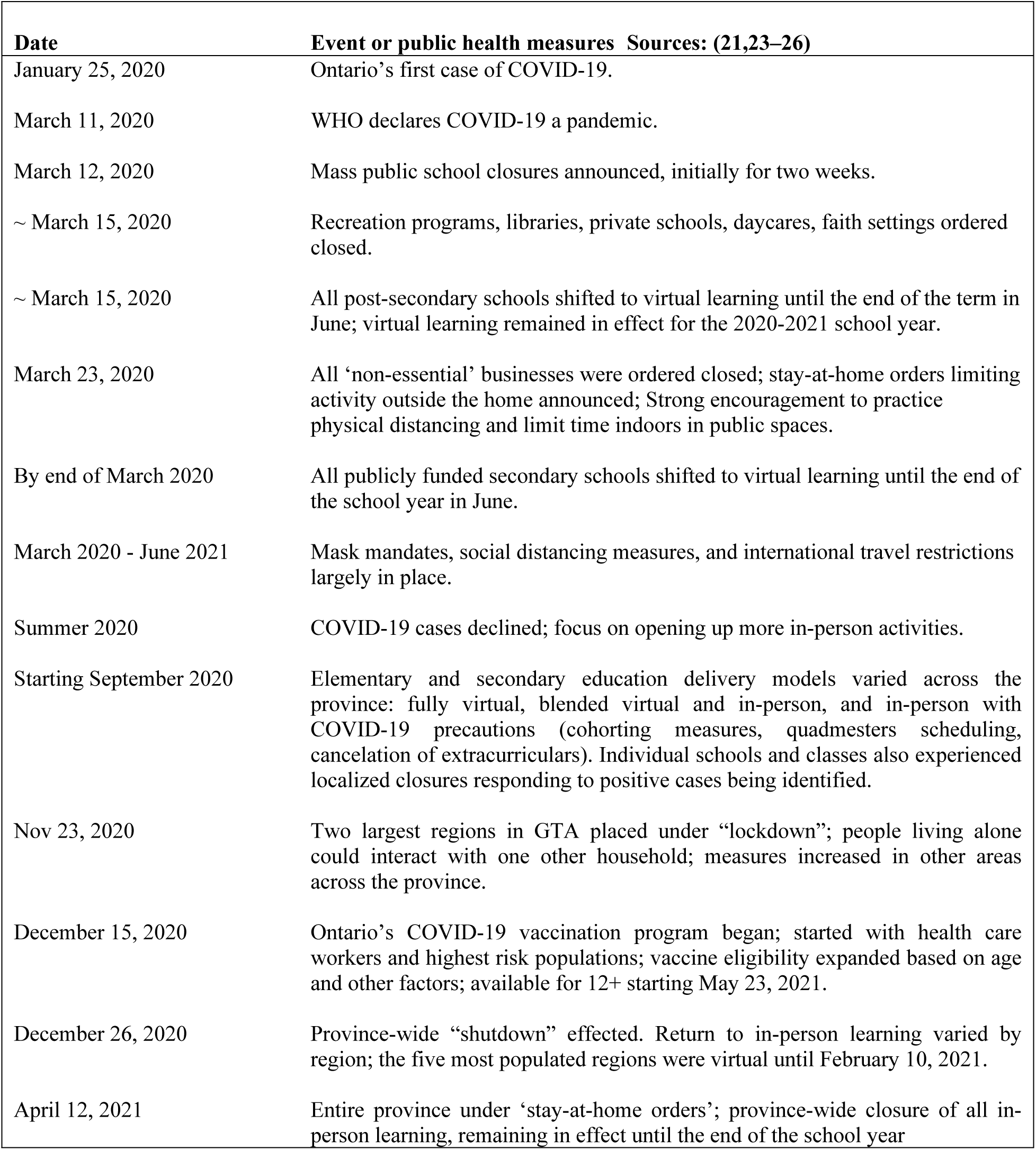
Ontario pandemic public health measures March 2020 – June 2021.

### Research question

We wished to explore how Ontario YYAs and the important foundations they build upon were impacted by the pandemic and related public health responses that had ebbed and flowed for over a year. Using qualitative semi-structured interviews we discussed related topics, the focus of this article. Interviews then turned to impacts on vaping decisions and behaviours, relevant to a broader research program this is a part of, funded by Canada’s Social Science and Humanities Research Council (SSHRC); findings on that will be reported elsewhere.

## Materials and methods

In reporting this qualitative research we have followed the consolidated criterial for reporting qualitative studies (COREQ) (27).

### Recruitment, respondents and interview methods

Research ethics approval was granted by Western University’s Research Ethics Board (REB). March 1, 2021 thru May 15 2021 participants were recruited through Facebook and Instagram posts advertising that research was being conducted “to understand how the pandemic has affected youth and young adults’ lives and activities”. 178 eligible respondents (ages 16-21, living in Ontario) completed a Qualtrics pre-screening questionnaire and provided contact information, demographic data and information about frequency of vaping, smoking and alcohol use (from not at all to daily). Purposeful sampling maximized diversity amongst respondents, e.g., in terms of age, gender, ethnicity (and vaping and smoking frequency). Those submitting the pre-screen were entered into a draw for one of four $40 retailer gift cards.

Due to public health pandemic restrictions interviews were conducted virtually over Zoom (n=18) or phone (n=1), during April to June 2021, conducted by two researchers (MG, SC). Average length was 32 minutes. Interviewers advised respondents to be aware of the degree of privacy where they were, that the researchers were alone, and that the interview could be stopped or questions skipped at any time. Because interviews were conducted virtually, with ERB approval verbal consent was obtained from participants; interviewers reviewed the Letter of Information (LOI) that had been emailed to invited participants in advance, checked the appropriate boxes for consent, signed the consent form, and emailed a scan of the fully executed LOI to the participant after their interview. With consent, interviews were audio/video recorded and verbatim transcripts produced and verified (n=18) or detailed notes taken (n=1). With a follow-up thank you e-mail, participants were provided contact information and links to age-appropriate mental health resources. Parental consent was not required or sought for this research.

Twenty respondents age 16-21 were interviewed; one was dropped from analysis due to responses that were highly inconsistent, leaving 19. Interviewers felt data saturation had been obtained at this point, meaning they were not hearing significantly new ideas in the last few interviews. Twelve respondents identified as female, seven as male (none self-identified otherwise). Eight were in high school, ten in post-secondary, and one worked full time. Nine self-identified as White, four as East, South or Southeast Asian, one as Latin American, one as Middle Eastern; and four as ‘mixed-race’; none identified as Black, First Nations, Metis, or Inuit. 14 lived in an urban/city area, four in suburban areas, and one rural. Each interviewed received a $30 retailer gift card.

A semi-structured interview questionnaire was pilot tested with two respondents; their interviews are included in analysis as only minor changes were made to the protocol following these. Consistent with growing recognition of the importance of including YYAs in research about YYAs (28,29) our research benefited from two university student researchers (MG and SC), one of whom was male, one female, who conducted interviews and were involved in analysis and manuscript preparation. Respondents were asked first to tell a bit about themselves, with prompts as needed for who they lived with, whether in school, employed, and how they liked to spend free time. Each was then asked how their life has been impacted by the pandemic and related restrictions, with follow-up prompts as needed to ask about impacts on relationships (with family, friends, significant others), school, employment, and important events. They were asked how concerned they were about getting COVID-19 and how they have been coping with all of the changes.

### Analysis Methods

Our approach was informed by reflexive thematic analysis (30–33), whereby we acknowledge that themes are not objective findings that anyone would reliably conclude ‘emerged from the data’, but instead are derived from researchers’ iterative engagement with, and subjective interpretation of verbal data. Like grounded theory approaches (34,35), reflexive thematic analysis requires iteratively engaging with the data, searching for patterns and shared meaning among responses, and developing themes that convey what the researchers take away given intense interaction with responses. Unlike grounded theory methods, thematic analysis does not require deriving testable theories that explain some phenomenon.

An initial reading of the transcripts and discussion about what we saw in the data was done by SC, MG, and LA who developed a preliminary set of concepts that we found compelling in the data. Three of the authors (MG, LA, CD) read the transcripts repeatedly to consider individual context and further develop themes. One of the authors (CD) brought experience and research expertise as a public health professional; two were themselves young adults (SC, MG); LA is a Behavioral Decision Theorist (PhD) with experience in mixed methods adolescent and adult decision making research. Transcripts were further analyzed by MG and LA using MaxQDA software to facilitate coding utterances that corresponded to topics of interest, facilitating our ability to identify similarities, differences, and patterns. The authors met several times to suggest, discuss, and refine themes. Participants did not provide feedback on the findings.

## Results

We identified the thirteen themes organized into four meta-themes: impacts on self; impacts on institutional foundations (education, employment, transitional events/rituals), impacts on relationships, and how respondents coped with all the changes. Clear in the coping theme is that mental health was a concern to respondents, arising purely in what respondents volunteered, as we asked no questions about mental health (we did ask how respondents coped with all of the changes). Tables 2-6 provide illustrative quotes for themes - we’ve placed those that exemplify two or more themes where we feel best offer unique insights.

**Table 2:**
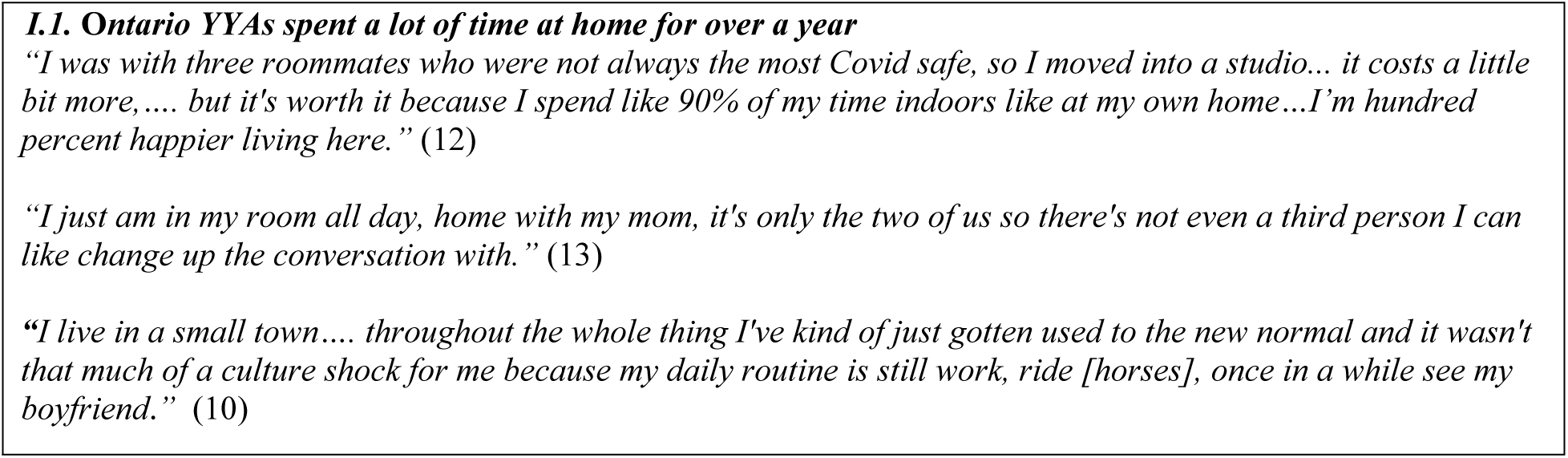
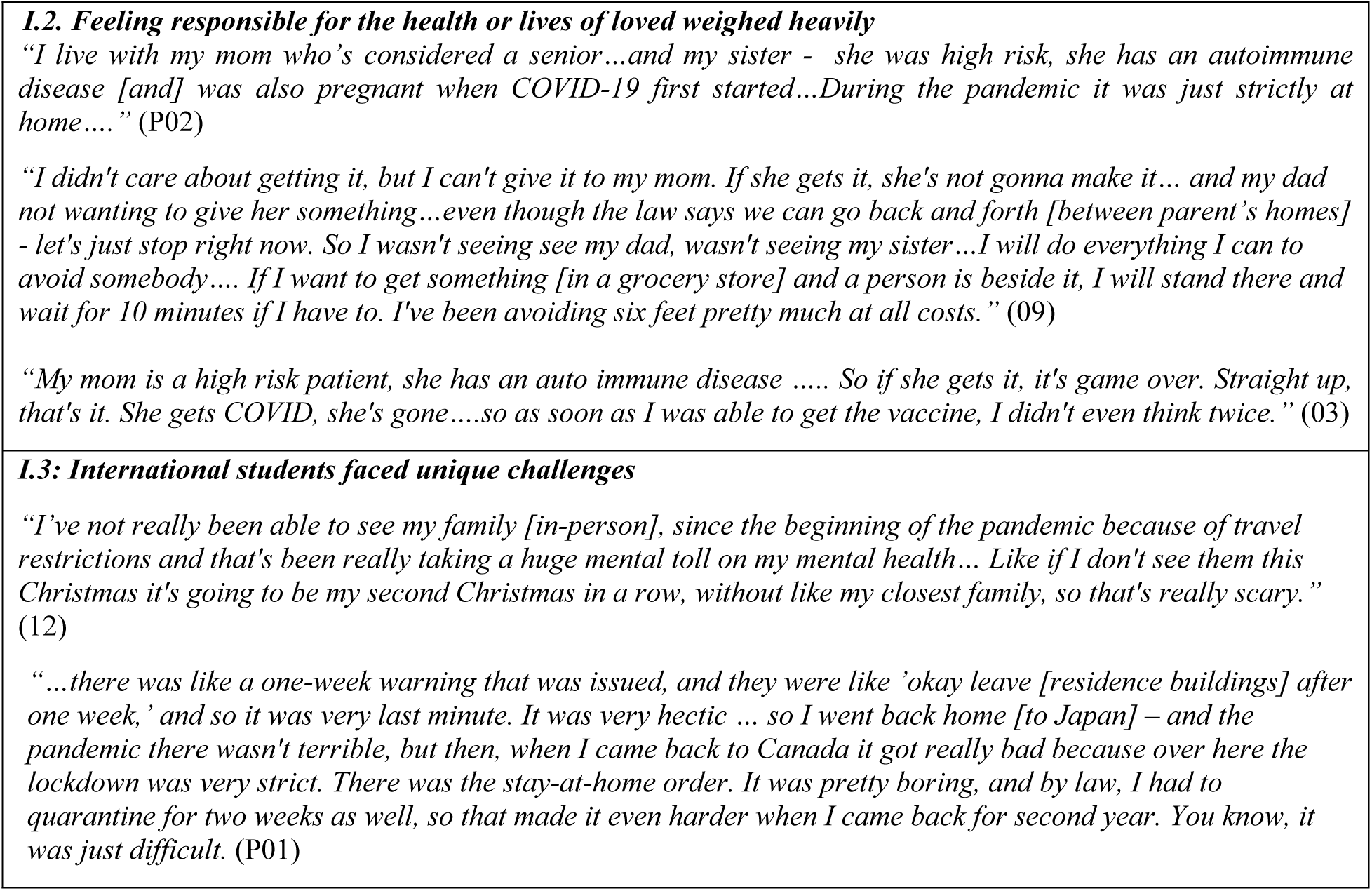
Impacts on self (excluding mental health)

### I. Impacts on Self

We identified three themes related to impacts on self: YYAs were at home a lot of the time; feeling responsible for the health and safety of loved ones weighed heavily; and, international students faced unique challenges.

#### I.1 Ontario YYAs spent a lot of time at home for over a year

One respondent who lived in a rural area and worked full time as an essential worker noted, “*even with all the lockdowns on and off I haven’t felt all that restricted*.” This sentiment was not shared by other respondents. These discussed how pre-pandemic they were seldom home, spending time at school, work, in extra-curriculars, and being out with friends, but during the pandemic were “always” at home. Some more specifically reported mostly being in the basement or their room. Some mentioned having returned to their parent’s home after leaving post-secondary residence early in the pandemic; one commented on having had to *“learn how to be a child again”*(*03*). A few referred to times during summer 2020 when the province ‘opened up’ and they got to live more normally. Illustrative quotes are found in Table 2.

Few seemed to question whether restrictions were necessary. Those who believed that peers attended parties or otherwise broke the rules (often based on social media posts) seemed perplexed or angry about such behaviour. One reported moveing away from roommates who did not follow public health guidance.

#### I.2 Feeling responsible for health or lives of loved ones weighed heavily

All participants expressed they were not very worried about getting COVID-19 themselves. Some noted times they had been worried, early in the pandemic or when someone they knew tested positive. Most volunteered why they were not worried: they were young, healthy, would only have flu-like symptoms, or would be able to recover if hospitalized. One noted that the new variant in spring 2021 was hospitalizing young people and so recently had been more concerned.

However, many discussed fears of passing COVID-19 to others (see Table 2 for quotes). Some did not want the *‘blame,’ ‘burden,’ ‘embarrassment,’* or to *‘have it on my conscious’* for spreading COVID-19 to classmates, teachers, parents, or siblings. Most living with family discussed family members they perceived as high risk for severe illness or death, perhaps suggesting ‘high risk’ equates to ‘highest risk in the family’, but with some extremely concerned about the high risk of fatal outcome if a specific loved one got COVID-19. Respondents described being “*worried*”, “*stressed*”, or “*terrified*” that high-risk loved ones might contract COVID-19, and about the possibility they might be the source of exposure. Some described going to great lengths to avoid others to protect loved ones; those seemed most likely to express relief offered by recently available vaccines.

#### I. 3: International students faced unique challenges

One respondent was an international post-secondary student from Japan; another was from Singapore. The first discussed the stress of having to suddenly leave Canada when residences were closed with little notice, then returning to experience a two-week quarantine in the fall. The second discussed the difficulties of having not seen family since before the pandemic. They moved to a small apartment by themself due to concerns about roommate (lack of) social distancing behaviours. Another student discussed being sad because international friends had left suddenly over a year earlier and not yet been able to return to Canada. See Table 2 for quotes.

### II. Impacts on institutional foundations

Impacts on school, employment, and important events were either volunteered by respondents, or discussed in response to prompts about these. We’ve organized these into four themes. Impacts on learning were many and varied, summarized into one theme on inferior learning models and motivation to learn. Respondents were concerned about impacts on professional development. Many were impacted negatively by changes in employment. Finally, loss of rituals and transitions was felt keenly.

#### II. 1: Many felt new learning models were inferior and decreased motivation

While a few respondents suggested learning was not negatively impacted, most reported it had been. A nursing student described how clinical training, typically in person, was now provided via virtual lectures. Respondents reported delayed support or less support than what they needed. Communication with, and access to, educators was challenging: teachers, guidance counselors, and professors were not as accessible or did not respond quickly to emails; presumably asynchronous email communication is not the same as being able to raise your hand during class with a question, immediate response, and dialogue. Illustrative quotes are found in Table 3.

**Table 3:**
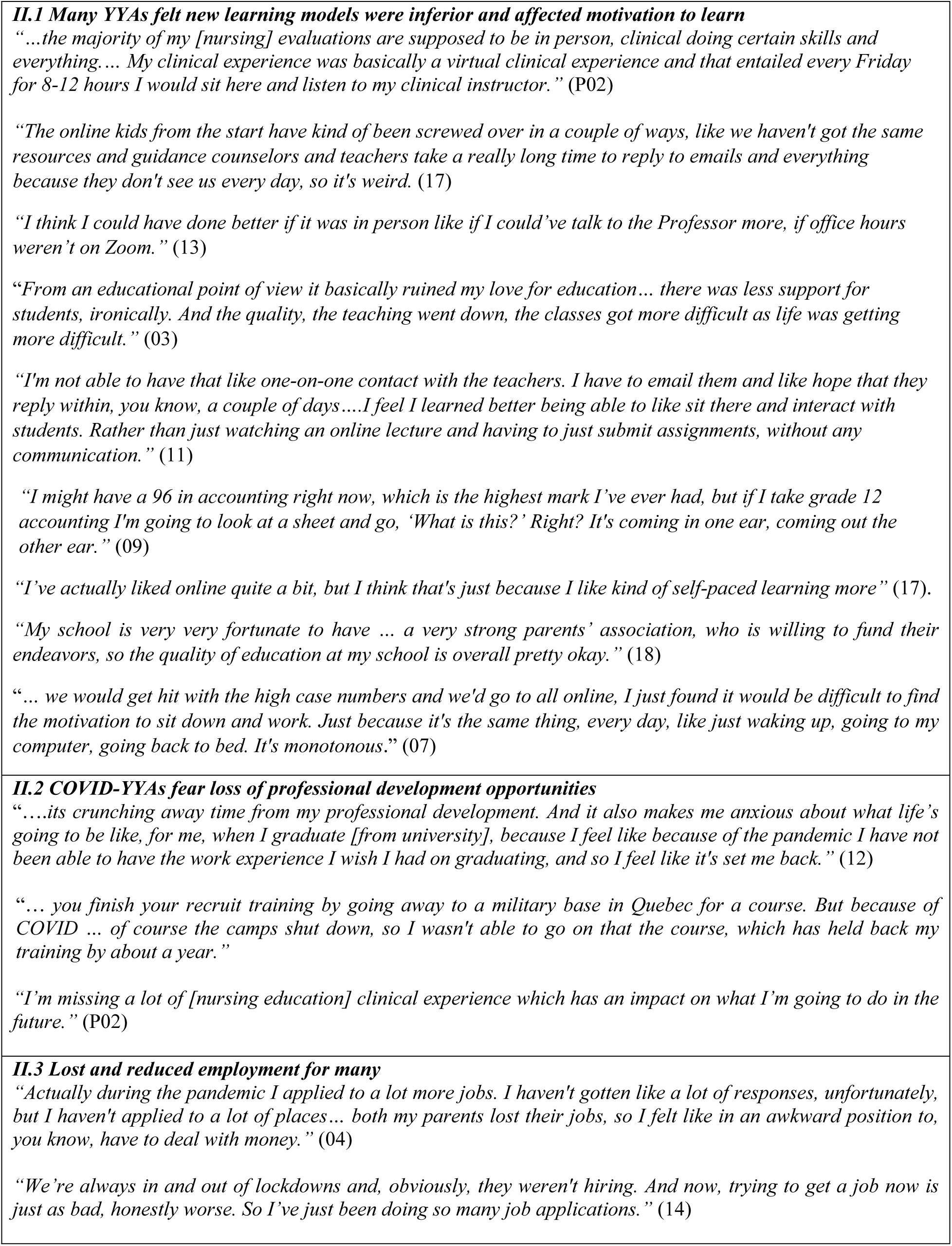
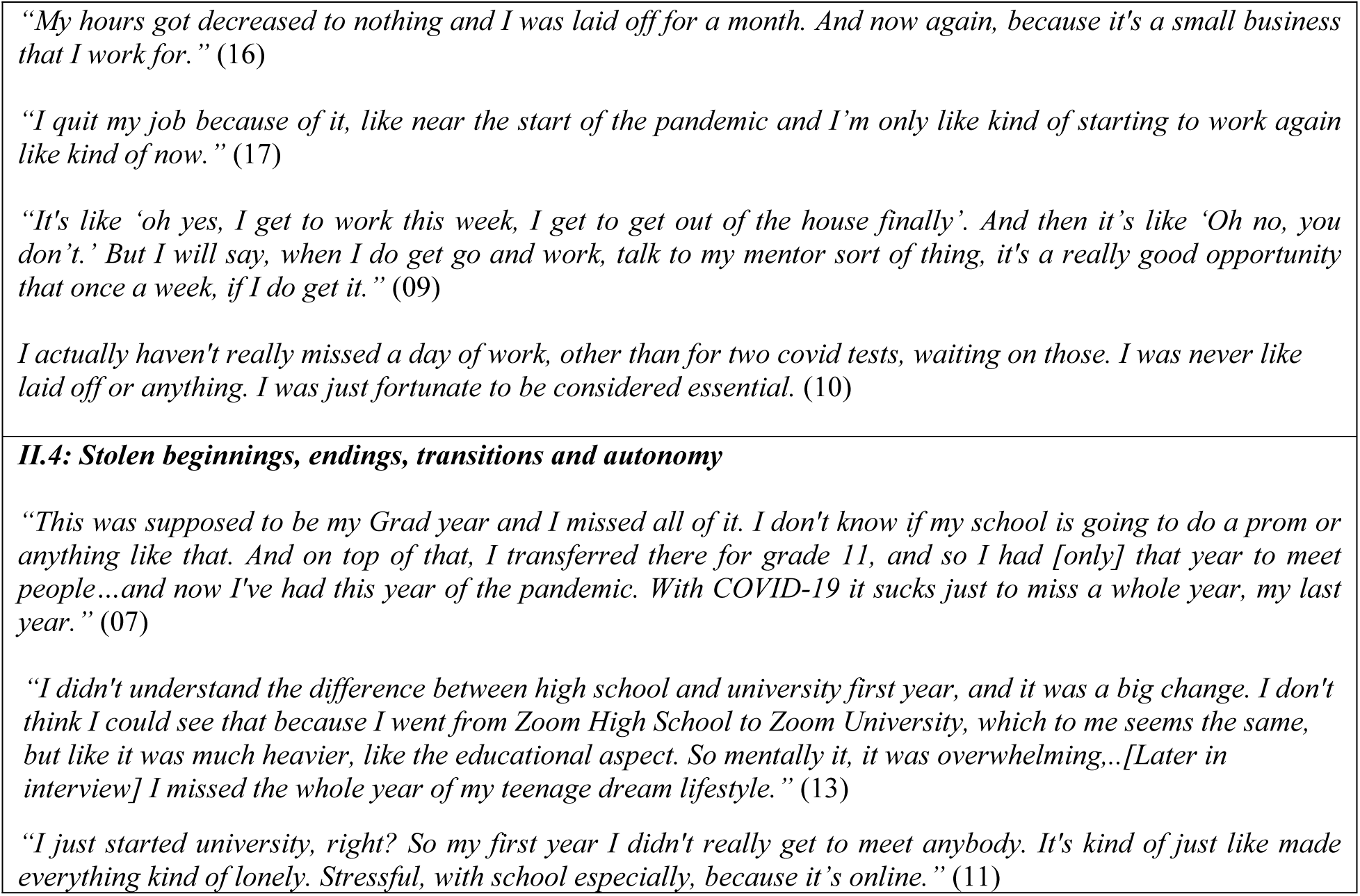
Impacts on institutional foundations youth build upon.

The sentiment *“Online school killed my motivation to learn,”* was not unusual. Many respondents felt they had learned less than they would have otherwise. One post-secondary student disliked the changes so much they got permission to rearrange the normal co-op schedule to avoid virtual learning. Some described physical impacts, such as eye-strain and migraines they attributed to too much screen-time with online school. We heard reports of lower quality instruction, inability to benefit from peer co-learning, and sub-par on-line assessment methods.

Students with Individual Education Plans (IEPs) were not always afforded their accommodations; for example, rather than a quiet space normally provided for exams, exams online at home were in the midst of distractions. Even students who had chosen online learning pre-pandemic, and who expressed that they preferred online learning, expressed frustrations with education changes brought by pandemic.

#### II. 2: COVID-YYAs fear loss of professional development opportunities

Some discussed how professional training and development opportunities had been negatively impacted, with concerns about long-term impacts on skills and careers (see Table 3 for illustrative quotes). A post-secondary student noted that while she was able to work in a co-op position, most of her friends had lost this opportunity due to employers suspending co-op programs. She felt “robbed” of her co-op experience because she had to work virtually from her bedroom. An engineering student was concerned about the lack of internship opportunities he believed were needed to secure a good job after graduation.

#### II.3 Lost and reduced employment for many

All participants were asked about employment status and whether employment had been impacted during the pandemic. Only two had never been employed; one was seeking employment (so far unsuccessfully); the other wanted to work to pay for further education but lived with immune-compromised grandparents and so could not. Of those currently working, most had had periods of reduced hours or unemployment during the pandemic (see Table 3). Many had lost or quit jobs due to the pandemic; some were currently seeking new opportunities, others had found new employment. A couple mentioned wanting to earn their own income because parents had become unemployed during the pandemic.

A common sentiment was that employment provided an opportunity to get out and interact face-to-face with others, although those who had transitioned to virtual work did not experience that. One felt fortunate to be an “essential worker” and had worked throughout the pandemic.

#### II. 4: Stolen beginnings, endings, transitions, and autonomy

Several discussed delayed or missed beginnings, endings, milestones, and transitions from one phase in life to the next (see quotes in Table 3). Those who were finishing grade 12 when interviewed (n=5) discussed losing their entire last year of high school, including important end-of-year events. One said that even though she might not have wanted to attend things like prom, she’d have liked having the choice. Three were completing their first year of post-secondary school when interviewed, meaning the pandemic struck at the end of their secondary school education. These discussed missing important end-of-secondary-school events and rituals like graduation, and because universities were virtual for the 2020-21 school year, they also lost out on important aspects of their first year of post-secondary education. This included, for example, including living on-campus, rituals that solidify the transition to post-secondary such as moving away from parents and participating in first year orientation activities, and meeting new friends.

### III. Impacts on relationships

We identified three sub-themes related to relationships with friends, parents, and dating: friend circles grew smaller and sometimes closer; relationships with parents varied over time; and, impacts on dating varied. A fourth theme on digital socializing and collaborating is also identified and discussed.

#### III.1 Friend circles grew smaller; some friends grew closer

Several participants reported they no longer engaged with acquaintances or peripheral friends. Some, especially in first-year post-secondary, were sad about missing out on the opportunity to meet new people and make new friends. Pain from not being able to be with friends was palpable. One mentioned their best friend’s family was still living in isolation, and they could only get together if physically distanced. However, several discussed becoming closer to some friends, or became closer to a small group of friends. Many discussed how their time spent in-person with friends had changed, meeting in smaller groups and often outdoors (e.g., walking, riding bikes). Those most likely to report seeing friends as they had before the pandemic were post-secondary students living independently, often with roommates. One reported getting together with friends on a regular basis *“in socially safe ways,”* but also acknowledged that there were times gatherings were not safe. A few mentioned that their boy/girlfriend was the only, or one of the few, people they spent time with. Illustrative quotes are in Table 4.

**Table 4:**
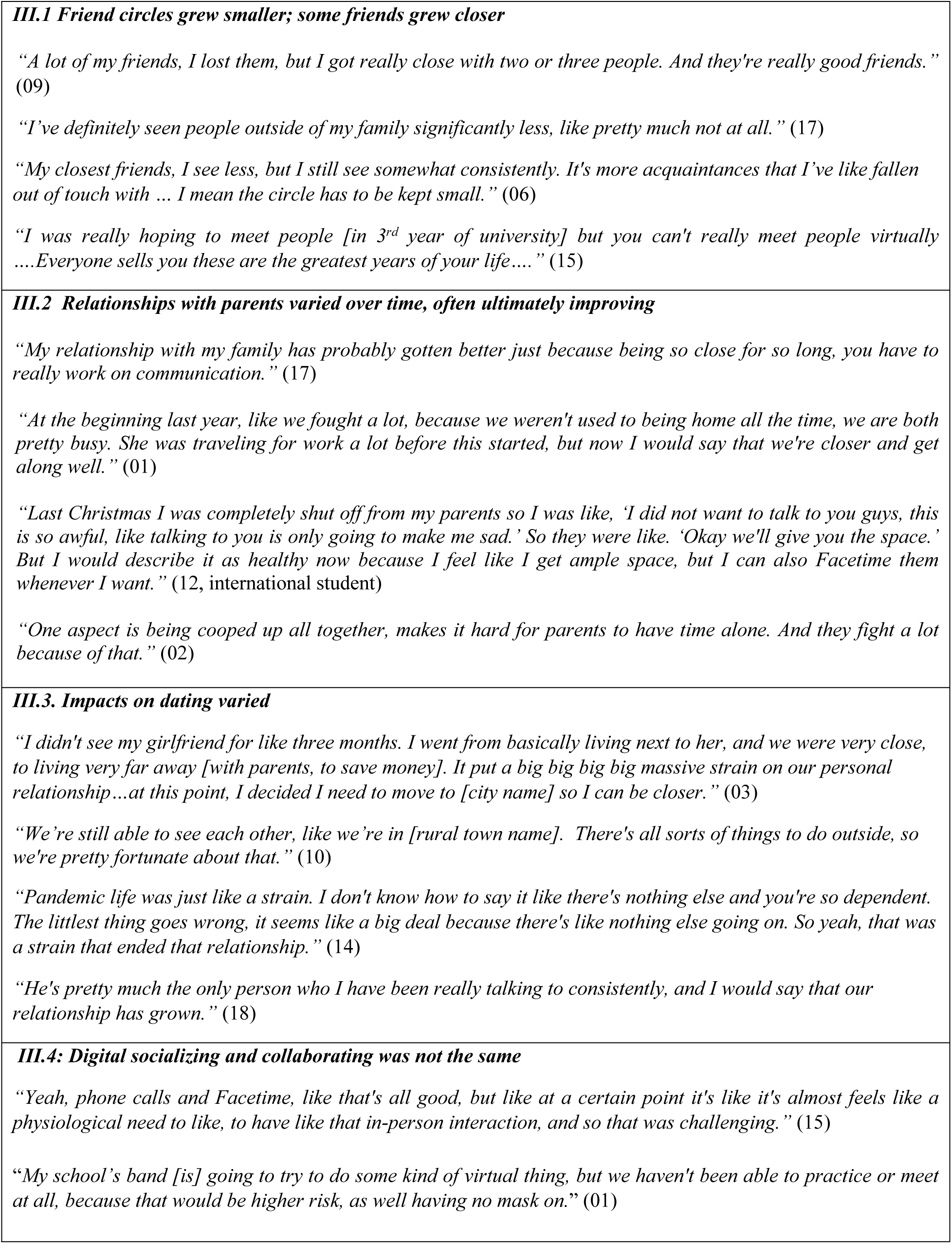
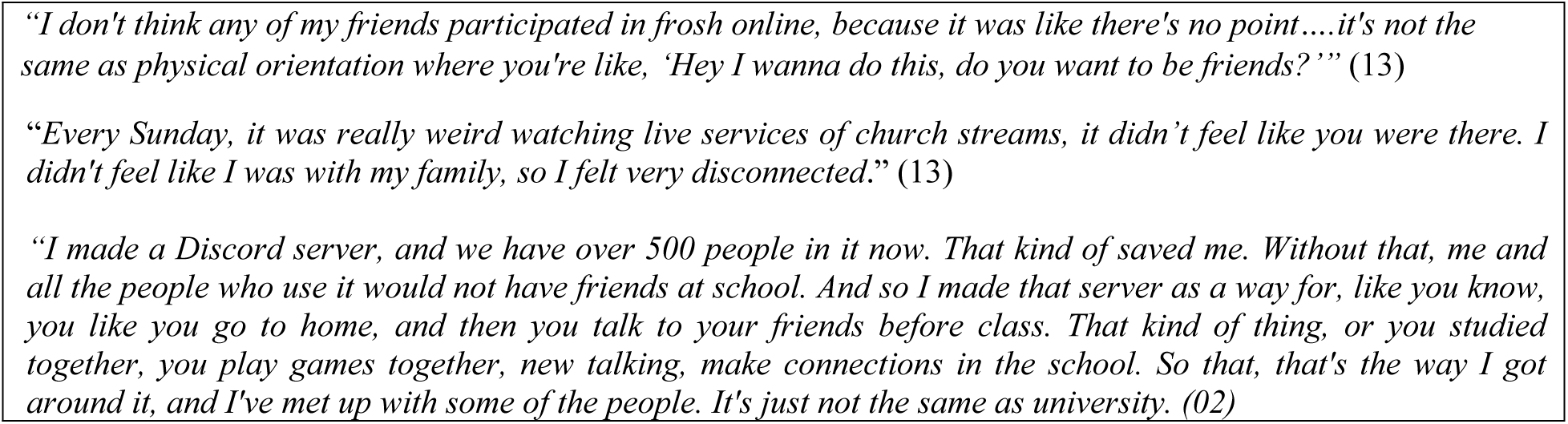
Impacts on relationships.

|  |
| --- |
| **III.1 Friend circles grew smaller; some friends grew closer**<br>*“A lot of my friends, I lost them, but I got really close with two or three people. And they're really good friends.” (09)*<br>*“I’ve definitely seen people outside of my family significantly less, like pretty much not at all.” (17)*<br>*“My closest friends, I see less, but I still see somewhat consistently. It's more acquaintances that I’ve like fallen out of touch with ... I mean the circle has to be kept small.” (06)*<br>*“I was really hoping to meet people [in 3<sup>rd</sup> year of university] but you can't really meet people virtually ....Everyone sells you these are the greatest years of your life....” (15)* |
| **III.2 Relationships with parents varied over time, often ultimately improving**<br>*“My relationship with my family has probably gotten better just because being so close for so long, you have to really work on communication.” (17)*<br>*“At the beginning last year, like we fought a lot, because we weren't used to being home all the time, we are both pretty busy. She was traveling for work a lot before this started, but now I would say that we're closer and get along well.” (01)*<br>*“Last Christmas I was completely shut off from my parents so I was like, ‘I did not want to talk to you guys, this is so awful, like talking to you is only going to make me sad.’ So they were like. ‘Okay we'll give you the space.’ But I would describe it as healthy now because I feel like I get ample space, but I can also Facetime them whenever I want.” (12, international student)*<br>*“One aspect is being cooped up all together, makes it hard for parents to have time alone. And they fight a lot because of that.” (02)* |
| **III.3. Impacts on dating varied**<br>*“I didn't see my girlfriend for like three months. I went from basically living next to her, and we were very close, to living very far away [with parents, to save money]. It put a big big big big massive strain on our personal relationship...at this point, I decided I need to move to [city name] so I can be closer.” (03)*<br>*“We’re still able to see each other, like we’re in [rural town name]. There's all sorts of things to do outside, so we're pretty fortunate about that.” (10)*<br>*“Pandemic life was just like a strain. I don't know how to say it like there's nothing else and you're so dependent. The littlest thing goes wrong, it seems like a big deal because there's like nothing else going on. So yeah, that was a strain that ended that relationship.” (14)*<br>*“He's pretty much the only person who I have been really talking to consistently, and I would say that our relationship has grown.” (18)* |
| **III.4: Digital socializing and collaborating was not the same**<br>*“Yeah, phone calls and Facetime, like that's all good, but like at a certain point it's like it's almost feels like a physiological need to like, to have like that in-person interaction, and so that was challenging.” (15)*<br>*“My school’s band [is] going to try to do some kind of virtual thing, but we haven't been able to practice or meet at all, because that would be higher risk, as well having no mask on.” (01)* |

*"I don't think any of my friends participated in frosh online, because it was like there's no point....it's not the same as physical orientation where you're like, 'Hey I wanna do this, do you want to be friends?'" (13)*
*"Every Sunday, it was really weird watching live services of church streams, it didn't feel like you were there. I didn't feel like I was with my family, so I felt very disconnected." (13)*
*"I made a Discord server, and we have over 500 people in it now. That kind of saved me. Without that, me and all the people who use it would not have friends at school. And so I made that server as a way for, like you know, you like you go to home, and then you talk to your friends before class. That kind of thing, or you studied together, you play games together, new talking, make connections in the school. So that, that's the way I got around it, and I've met up with some of the people. It's just not the same as university. (02)*

#### III.2 Relationships with parents varied over time, often ultimately improving

Many participants reported improved relationships with parents during the pandemic. Several said that relationships were strained early in the pandemic, but had improved since. Some discussed ongoing issues, not so much in their relationship with parents, but in their parents’ relationship with each other. Qualitative research early in the pandemic found adolescents reported struggles with family conflict, and stressed and unsupportive family members (Scott et al., 2021); the current study is suggestive that after a more extended periods with stay-at-home orders, these relationships actually improved. See Table 4 for illustrative quotes.

#### III. 3: Impacts on dating varied

About half the respondents discussed dating or relationships with significant others. Several discussed how the pandemic impacted dating, for example having to move back to a parent’s home created physical distance and stress; trying to maintain a relationship via Face-time; only seeing each other out-of-doors; the fact that things were easily blown out of proportion as pandemic stresses added up. We heard of delaying a first romance because of inability to be together in person. Some discussed breaking up, while others started a relationship during the pandemic; such events of course may have had nothing to do with the pandemic. At least one respondent noted they had no out-of-pandemic experience to compare starting or ending a relationship to. See Table 4 for illustrative quotes.

#### III.4: Digital socializing and collaborating is not the same

Respondents reported that as with learning, socializing had shifted to virtual platforms. They talked about face-time with friends, “*maintaining friends*” online, socializing virtually via Netflix and Discord parties, playing video games, and virtually consuming substances together. For some, work moved online. School and extra-curricular activities such as band, dance competitions, and graduation ceremonies became virtual activities. While time together digitally helped, it was not the same as being in person with people. See Table 4 for illustrative quotes.

### IV. How respondents coped with all the changes

Although we did not ask about mental health, many participants volunteered comments throughout their interviews about stress, anxiety, mental health, being lonely, finding things hard, etc., summarized in the theme, *This all took a toll*. We did ask respondents how they had coped with all the changes, and found diverse coping strategies as summarized in the second theme, *Coping strategies were diverse*.

#### IV.1 This all took a toll, including on mental health

We heard of “*monotonous*” “*repetitive*” days that “*melted together*”, of being lonely and unable to see friends. Several used the term “*disconnected*.” We heard of having felt overwhelmed, losing motivation to learn, socialize, or leave the house. Respondents discussed feeling stressed, lack of control, the unknown of when it will all change, when it will feel ‘normal’ again, being diagnosed with depression or anxiety, or worsening mental health conditions. Often this came up when discussing something that had been lost as conveyed in many of the themes above. One expressed, “*After a certain point, like just, like you just have to start laughing at it, you know, because it’s like if you don’t laugh at it, like you’ll just cry about it. So like, which one do you want to do”* (15, 3rd year post-secondary). See other illustrative quotes in Table 5.

**Table 5:**
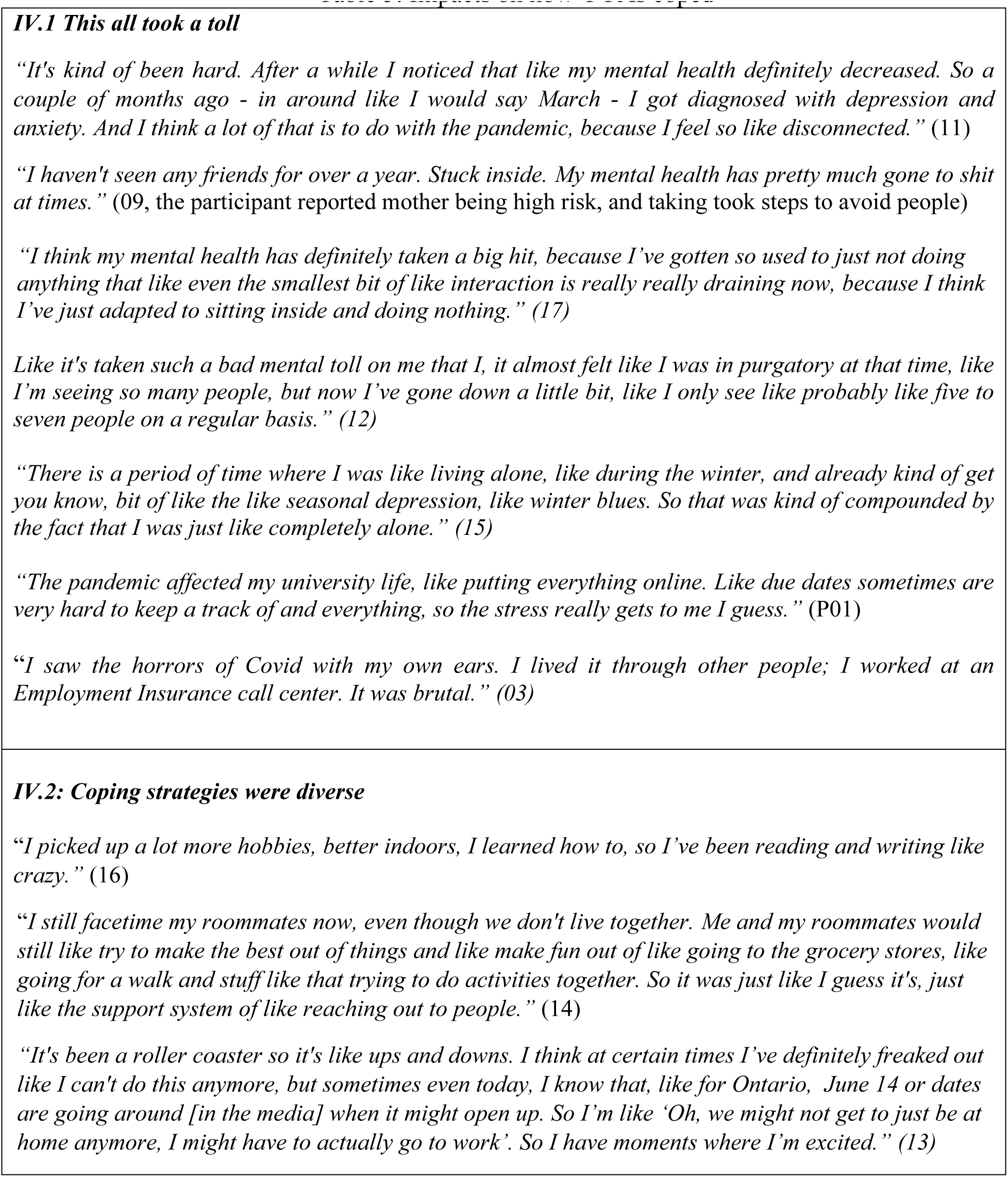
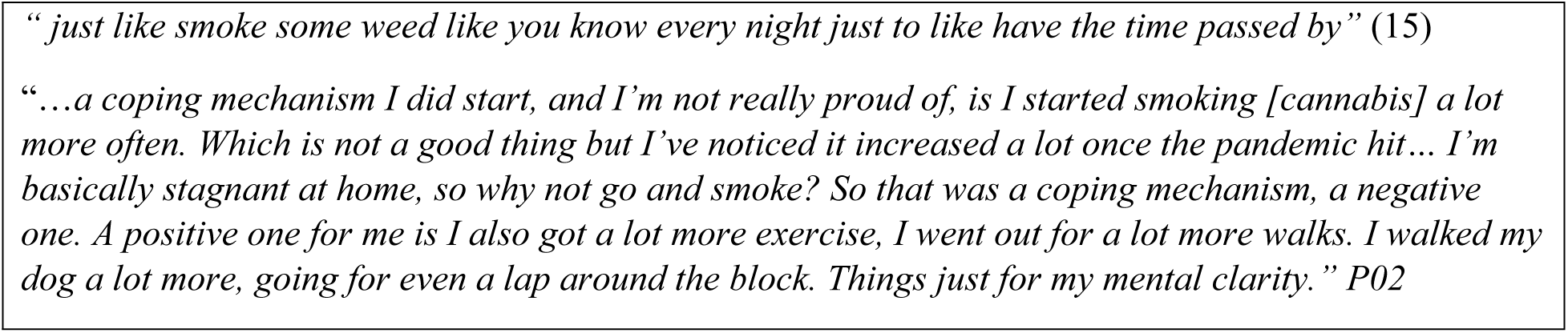
Impacts on how YYAs coped.

#### IV.2: Coping strategies were diverse

We asked participants how they were coping with all of the changes. Some volunteered they smoked, vaped, or used cannabis to cope or relieve stress (see example quotes in Table 5). Some discussed going for walks with friends, discovering new activities/hobbies, exercising more. One was proud of having started a discord server to allow students to socialize and work together.

Coping strategies may have shifted over time. Some discussed looking forward to when the restrictions would end. One secondary school student who’s mother was at high risk and who reported generally not seeing anyone described what he’d done to cope:

“*This semester, I kind of started to put a thing in place and go: okay I’m going to use excel and I’m going to make a schedule, I’m going to look at this. You know, I’ve done too much work, I’m going to go outside, go for a bike ride. Setting myself goals. So I’m trying to hit 10K in 30 minutes, which I hit the other day, and it was the first time I felt euphoric in a long time.”* (09)

## Discussion

The present study sought to explore how youth and young adults in Ontario Canada were impacted by over a year of the COVID-19 pandemic and public health responses to limit viral spread through analysis of rich verbal data. We’ve shared thirteen themes we took from the interviews, several of which reflect loss. Some losses were the result of not experiencing something that was looked forward to, such as important rituals, moving away from home, meeting new friends in a next phase of life, a first job, or first romance. Some losses represented loss of something in hand, such as time with friends, collaborative learning, and employment. Other loses represented lost potential, in terms of knowledge learned and skills needed for future success and well-being. Respondents discussed measures they took to protect themselves, for example, moving back to parent’s home, which also led to losses, for example being physically distanced from friends. The losses were deeply felt. And yet, our sense was that our respondents largely did not question the necessity of public health measures; many expressed negative feelings towards others who they perceived were not following the rules.

As found in studies early in pandemic, participants discussed loved ones believed to be at high risk of COVID-19 and their need to protect them (13,14,36). There was some sense that ‘high risk’ meant ‘highest risk in household’, and that feeling responsible for loved ones’ health was fairly common. However, some had family members who they believed were likely to suffer a fatality if they contracted COVID-19. These youth were especially likely to discuss extra precautions they took, delaying employment, avoiding physical proximity to people, and seldom seeing friends, to protect loved ones. YYAs who felt responsibility and accountability for loved ones’ health seemed impacted by the stress of having to constantly weigh their own and others’ risk in decisions to leave home or see friends, extended over more than a year. These youth seemed especially likely to express relief that COIVD-19 vaccinations, still a recent innovation, were now available. The extended period of stress and worry could have long term impacts; this cannot be known without further research.

A subset of our participants experienced loss and unhappiness over delayed or missed milestones. Prominent was unhappiness around missing the end of secondary school in spring 2020 or spring 2021. Feelings expressed are consistent with findings from an extensive stream of research showing that “people’s global evaluations of past affective episodes can be well predicted by the affect experienced during just two moments: the moment of peak affect intensity and the ending” (37,38). In other words, how something, e.g., the last year of secondary school, will be remembered will be based on feelings at the low or high point during the experience, or feelings as the experience ends. This stream of research shows that when people approach the end of something important to them, they tend to spend more time with close friends. This was not possible given pandemic response restrictions, perhaps adding to the intensity of the loss.

Like studies early and others later in the pandemic, our respondents discussed lost milestones such as graduation associated with learning (13,14,18). Our respondents who missed secondary school graduation in 2020 all began post-secondary education that fall. They were not able meet other students, faculty, or staff face-to-face, and are unique in Canadian (and elsewhere) history for starting post-secondary education completely virtually, not by choice, and with virtual learning models that were new to all involved and evolving in real-time. Our sense is that these young adults were especially impacted by the pandemic, due to the loss of important endings *and* not being able to transition to important new beginnings as expected, for example, leaving home and experiencing new approaches to education. Its’s important to note that a norm in many Canadian universities is for students to live in residence on campus only in first year and then move off-campus; missing this, and other opportunities to form close friendships in first year, could have lasting impacts that can only be assessed with further research. Our sample is limited in lacking secondary school respondents who did not start/plan further education, and so we cannot comment on the population that went directly into employment (or unemployment).

All of the changes meant not seeing friends in-person, having fun, hanging out, learning together. This was keenly felt by our respondents who expressed feeling lonely, missing their friends, and having mental health impacted, like findings in a large body of research that has examined mental health of YYAs during the pandemic (e.g., Khan et al., 2020; López-Castro et al., 2021; McCluskey et al., 2021). We heard from respondents that new virtual learning models and changes to in-person schooling, hastily introduced at the start of the pandemic and evolved over the next year, did not measure up to traditional learning models. Respondents told us of monotonous days, largely alone, at home. Many volunteered that they had been lonely, stressed, anxious, or depressed, and that their mental health had been impacted. While we did intend to study mental health, and do not claim to have assessed it in any way, clearly their own mental health was on the minds of many of our respondents. Given the extended periods of stay-at-home orders in Ontario, and high number of days spent in virtual learning, we would be especially concerned for lasting impacts on some YYAs in Ontario and other localities with extended public health measures.

Perhaps a bright spot in the pandemic was that some relationships deepened and improved for some YYAs during the extended pandemic. Several discussed difficult relationships with parents early in pandemic, as reported in other studies from early in pandemic (13,14). However, several went on to report that their relationships evolved, leading them to feel closer to parents at the time interviewed. We also heard from several respondents that they developed closer relationships with some friends during the pandemic.

## Conclusions

There was only a short window during which research on the impacts of the COVID-19 pandemic could be conducted. Our study is one of few qualitative studies published on impacts to YYAs after over a year of pandemic, taking place in Ontario Canada, which, as noted earlier, had ‘stay-at-home’ orders in place over much of that year, the Canadian province with the most weeks of in-person school closures. While our sample was small, it is suggestive that YYAs who lived with family members perceived to be at highest health risks from COVID-19, and those who were in their final year of secondary school in spring 2020, entering post-secondary in fall 2020, suffered unique impacts that could have long-lasting effects.

While our research is limited by a small sample, discussions were rich and illuminating, and sometimes evoked strong emotions. The YYA population is very diverse and not all would have had the same experiences. While some really struggled, and some reported turning to substances to help cope, others were able to cope and saw improvements in close relationships. This study provides some insights into YYA behaviour and adapted coping strategies employed during an extended pandemic period of high uncertainty and change.

## Data Availability

This project received ethics approval from Western University Research Ethics Board (REB) We did not seek consent to publicly share anonymised interview transcripts. REB approval does not extend to publicly sharing data beyond the research team who meet the criteria for access to personal data. The ERB approved Letter of Information provided to respondents, and which was the basis of their consent to participate, stated, "By consenting to participate in this study, you are agreeing that your data can be used by the research team for new research purposes (e.g., to answer a new research question). That future research may be in collaboration with other researchers." Researchers interested in such collaboration should contact the corresponding author.

## Footnotes

1 Provincial terminology and definitions for periods with the most restrictive measures changed over time, including use of the term ‘stay-at-home’; colloquially these were generally referred to as ‘lockdown’.

